# The influence of fetal sex on maternal blood pressure in pregnancy

**DOI:** 10.1101/2025.01.28.25321287

**Authors:** Caitlin S. Decina, Robin N. Beaumont, Julius Juodakis, Nicole M. Warrington, Kashyap A. Patel, Pål R. Njølstad, Stefan Johansson, Andrew T. Hattersley, Bo Jacobsson, William L. Lowe, David M. Evans, Rachel M. Freathy

## Abstract

**Background:** Pregnancy with a male fetus carries a higher risk of term pre-eclampsia than pregnancy with a female fetus. Based on evidence that maternal blood pressure (BP) may be raised in pregnancies with Beckwith-Wiedemann syndrome (fetal overgrowth), a possible contributing factor to the association between male sex and term pre-eclampsia is that males grow faster, reaching ∼130 g higher birth weight, on average, than females. The association between fetal sex and maternal BP in healthy pregnancies is not known. We hypothesized that male sex would be associated with higher maternal BP in healthy pregnancies, and that this association would be explained by birth weight differences between males and females.

**Methods and findings:** We tested the association between fetal sex and maternal systolic (SBP) and diastolic blood pressure (DBP), measured at ∼28 weeks of gestation, in a meta-analysis of five different cohorts of mother-child pairs (n up to 109,842). Maternal BP was analyzed as both a continuous and dichotomized (high BP: yes or no) outcome. Linear regression models were constructed with and without adjustment for birth weight to assess whether any difference in maternal BP was explained by the difference in birth weight between male and female babies. Lastly, we constructed a fetal genetic score for birth weight using 186 own-birth-weight-associated single-nucleotide polymorphisms (SNPs) to test whether birth-weight-raising-alleles in the fetus were associated with maternal BP in pregnancy (n up to 32,232). Both maternal SBP and DBP were higher in pregnancy when carrying a male fetus compared to a female fetus (mean difference 0.35 mmHg [95%CI: 0.15-0.55] and 0.35 mmHg [95%CI: 0.21-0.49], for SBP and DBP, respectively). An independent effect of fetal sex remained when including birth weight but attenuated slightly (0.22 mmHg [95%CI: 0.02-0.42] and 0.31 mmHg [95%CI: 0.17-0.45], for SBP and DBP respectively). A positive effect estimate was found for odds of experiencing high maternal BP given pregnancy with a male fetus, but confidence intervals were wide (OR 1.05 [95%CI: 0.98-1.12]). No evidence for an association was found between a fetal birth weight genetic score and SBP or DBP when conditioned on maternal genotype.

**Conclusions:** We found strong evidence to support a small effect of male fetal sex on higher maternal BP in pregnancy and that larger fetal size at birth does not contribute to a substantial part of this association. Our findings do not indicate a difference in maternal BP that would warrant changes to routine monitoring in clinical practice but do suggest that male sex may be a contributing risk factor for BP-related complications.

## Introduction

Much of human pregnancy-related research investigates the effects of maternal physiology on the development of the fetus. However, the fetus may also influence maternal physiology. There is evidence to suggest that expectant mothers are at greater risk of experiencing different pregnancy complications depending on whether they are carrying a male or female fetus. In the literature, a higher risk of stillbirth^1^, preterm birth^2–5^, gestational diabetes^6^, and hypertensive complications such as term pre-eclampsia^6^ is reported for mothers carrying male babies, compared to those carrying female babies. Altered mechanisms of placental function may underlie these differences with males more susceptible to abnormal placentation and impaired responses to stress^7,8^.

Another possible reason for the observed sex differences is that male babies are larger and demonstrate faster fetal growth than their female counterparts^9,10^. One example of greater fetal growth leading to maternal clinical outcomes is when the fetus has Beckwith-Wiedemann syndrome, where mothers carrying macrosomic babies develop gestational hypertension at a higher rate compared with normal sibling controls^11^. Cases such as these have led to the idea that variation in the fetal genome can influence maternal physiology to enhance nutrient transmission and optimise growth. This idea is known as the fetal drive hypothesis, which posits that the genotype of the offspring may drive, at least in part, the physiology and behaviour of the mother^12,13^.

Genetic studies have predominantly investigated the effects of maternal genotype and physiology on the development of the fetus^14,15^, but less so potential effects of fetal genetics on maternal physiology in pregnancy. A previous study that explored this issue found that fetal alleles that increase birth weight are associated with a shorter gestational duration, and a higher blood pressure (BP) in the mother^16^.

Given the observed links between higher birth weight and raised maternal BP, we hypothesized that male sex would be associated with higher maternal BP in large datasets of healthy pregnancies, and that this association would be explained by birth weight differences between males and females. We aimed to investigate (i) whether mothers carrying male babies (on average ∼130 g heavier at birth^3,17^) have higher BP during pregnancy than those carrying female babies, (ii) whether any observed differences in maternal BP were driven by the difference between male and female birth weight, and (iii) whether a higher fetal genetic score for birth weight, conditional on maternal genotype, was associated with maternal BP.

## Methods

We tested the association between fetal sex and maternal BP, measured at approximately 28 weeks of gestation, within five different cohorts of mother-child pairs mainly of European descent, and meta-analysed the results across cohorts. Analyses were run in models where maternal BP was measured as a continuous trait, as well as in models where maternal BP was dichotomized (high BP: yes or no). Models were constructed with and without the inclusion of fetal birth weight to assess whether any difference in maternal BP was mediated by the difference in birth weight between male and female babies. Lastly, we constructed a fetal genetic score for birth weight to test whether birth-weight-raising-alleles were associated with maternal BP in pregnancy.

### Study populations

#### Hyperglycemia and Adverse Pregnancy Outcome (HAPO) Study

The Hyperglycemia and Adverse Pregnancy Outcome Study is an observational study which sought to determine the risks of adverse outcomes associated with varying levels of elevated maternal glucose that do not constitute overt diabetes mellitus^18^. Participants underwent a standard oral glucose tolerance test (OGTT) using a 75 g glucose bolus administered between 24 and 32 weeks of pregnancy, with 28 weeks being the targeted time for testing^18^. Of the eligible women across 15 centres in nine countries, 28,562 agreed to participate, 25,505 of whom completed the OGTT, leaving 23,316 available for analysis following further filtering^18^.

At the time of the OGTT visit, women’s height, weight, and BP were also measured^18^. Neonatal anthropometrics, including birth weight, and gestational age were recorded within 72 hours of delivery using standardized methods across all centres^19^. Genetic samples were obtained through blood collected from participating women at the time of glucose testing, while newborn DNA was isolated from umbilical cord blood samples taken at the time of delivery^20^.

A total of 2000 mothers and babies (1000 mother-child pairs) were genotyped using the Illumina Infinium^TM^ Global Screening Array. Quality control (QC) was performed where individuals with genotype call rate <97.5% (82 samples) were removed and SNPs removed if they had call rates <98% (2.9% of SNPs) or had Hardy-Weinberg equilibrium (HWE) P <1×10^−6^ (0.05% of SNPs). Genetically determined sex was compared with phenotypically determined sex, excluding samples where conflicts were identified (17 samples). Relationships for mother-offspring pairs were confirmed using KING software^21^, applying default KING thresholds for kinship, with samples excluded where the expected relationship did not match the reported relationship (14 samples). To derive the genetic ancestry of individuals, principal components (PC) analysis was performed on the genotype data using flashPCA^22^. Visual inspection of PC plots identified outlying individuals for exclusion who did not fit within an ancestry PC cluster (155 samples removed).

Upon completion of QC, a total of 1,732 (including 565 complete mother-child pairs) individuals and 671,798 SNPs were available for further genetic association analysis. Samples were phased using SHAPEIT2^23^, using the duohmm option^24^ to make use of known family relationships. Imputation was carried out using the Michigan Imputation Server^25,26^ with samples imputed to the HRC v1.1 reference panel^27^.

#### Avon Longitudinal Study of Parents and Children (ALSPAC)

The Avon Longitudinal Study of Parents and Children (ALSPAC) is a prospective observational cohort study. All pregnant women living in a defined area of Avon, UK expected to have a live birth between 1^st^ April 1991 and 31^st^ December 1992 were eligible for participation. Initial recruitment gave rise to 14,541 pregnancies of which there were 14,676 known fetuses, resulting in 14,062 live-born children. Among those live births, 13,988 were alive at 1 year of age^28,29^.

Maternal BP measurements were taken as part of routine clinical care during pregnancy by a health professional, and later abstracted from obstetric records by six trained research midwives^30^. Error rates were consistently <1% in repeated data entry checks, with no variation between midwives in mean values of abstracted data^30^. Each BP measure was taken once for women in the seated position using the appropriate cuff size, and measuring DBP using Korotkoff phase V^30^. Maternal age at delivery, offspring’s sex and birth weight in grams were also obtained from obstetric records^30^. Questionnaires administered to each woman during pregnancy were used to obtain further anthropometric measures and lifestyle information including maternal height and weight before pregnancy, parity, education, smoking status, and ethnicity^30^. Note that the study website contains details of all the data that is available through a fully searchable data dictionary and variable search tool (http://www.bristol.ac.uk/alspac/researchers/our-data/).

Repeated blood sampling was performed for each woman during routine antenatal care in addition to further sampling during follow-up clinics for both mothers and children, allowing for the collection of DNA samples for 11,343 children and 10,321 mothers^28,29^. Genotyping of maternal genetic data was performed using the Illumina 660 Quad Array and fetal data was genotyped using the Illumina 550 Quad Array. Genotype data were then imputed against the HRC v1.1 reference panel^27^ using the Michigan imputation server^25,26^ following QC: exclusion based on MAF <1%, HWE P<1×10^-6^, sex mismatch, kinship errors, and 4.56 SD from the cluster mean of any sub-populations cluster^31^. Ancestry PCs were generated in the 1000 Genomes sample in order to classify individuals according to the 3 main 1000 Genomes populations (European, African, South Asian) and separate out those of European genetic similarity^31^.

For all association analyses in the current study, we retained births which were deemed “survivors” under Wigglesworth classification^32^. For the associations between fetal sex and maternal BP, we restricted to self-reported White European ethnicity due to lack of power in the other available ethnic groups, and for the genetic score associations with maternal BP, restricted to those of genetically determined European ancestry (a subset of the self-reported White European ethnicity group). As not all individuals in the study had a DNA sample, self-reported ethnicity was used for the phenotypic analyses to maximize the sample size for association testing that did not rely on genetic data.

#### Born in Bradford (BiB)

Born in Bradford (BiB) is a multi-ethnic cohort study established in 2007 in Bradford, UK, including individuals of White European ancestry, and South Asian ancestry of largely Pakistani origin and a smaller sample of Bangladeshi origin^33^. Women eligible for participation in the study were those booked in for delivery at the Bradford Royal Infirmary and offered an OGTT at 26-28 weeks gestation^33^. Between March 2007 and November 2010, >80% of women who attended the OGTT accepted the invitation to participate, thus recruiting 12,453 women with 13,776 pregnancies^33^. Upon recruitment, women completed a comprehensive interviewer-administered questionnaire as well as being weighed, measured, and blood samples collected. Infants had anthropometry assessed at birth and blood collected via umbilical cord samples^33^.

Participants within the BiB study had BP measured multiple times over the course of pregnancy, with numerous measures taken within the same gestational week for some women. For our analyses, we sought to determine one BP measure taken as close to 28 weeks as possible for each unrelated woman (up to third degree relatives excluded based on kinship analysis) included in our sample to be consistent with BP data available in the other included cohorts for which BP was measured only once at 28 weeks. We devised a protocol to take the singular BP measure at 28 weeks if only one was present, otherwise, to take the median of multiple measures at 28 weeks. If measures at 28 weeks were not available, we expanded to a range of two weeks either side of 28 weeks (26-30 weeks) to maximize sample size, with a stepwise approach to then take the measures at 27 and 29 weeks together and select the median; if measures at those time points were unavailable, to then take the median of the measures at 26 and 30 weeks.

In our stepwise approach of BP measure selection, observations within each pregnancy were first sorted by SBP so that SBP was ordered lowest to highest over the range of time. Where there were instances of multiple BP measures to choose from, if the number of measures was odd, we took the middle (median) observation to use in analysis as mentioned above. If the number of measures was even, we looked to gestational age to decide which of the two middle values to use and took the later measure (latest GA). For instances where there were two BP measures on the same gestational day, the first measure was used.

A total of around 20,000 individuals were genotyped from collected biological samples. Both maternal and fetal data were genotyped using two separate array chips, Illumina HumanCoreExome and Illumina Infinium Global Screening (GSA)^34^. Individuals were excluded based on mismatch between genotypically-derived sex and documented phenotypic sex (n = 110), being genetic duplicates (n = 127), or mismatch among reported first-degree relative relationships (n = 470), comprising 707 total exclusions, approximately 3.5% of the original sample. PC analysis was performed using flashPCA^22^ software, using the first two PCs to classify ancestry as either White European or South Asian. No exclusions based on relatedness were made.

Genotype imputation was subsequently performed on 7,606 genetically defined Europeans and 9,248 genetically defined South Asians (n = 8,692 Pakistani and n = 556 Bangladeshi), with individuals of Pakistani and Bangladeshi ancestry imputed separately. Imputation was performed on each chip separately as the number of variants overlapping both chips was low, resulting in sample sizes of n = 5,707 Europeans (2,405 children, 3,302 mothers), n = 6,919 Pakistani (3,215 children, 3,704 mothers), and n = 424 Bangladeshi for the CoreExome chip (197 children, 227 mothers), and n = 1,899 European (614 children, 728 mothers), n = 1,773 Pakistani (784 children, 704 mothers), n = 132 Bangladeshi (54 children, 64 mothers) for the GSA chip. Prior to imputation, variant filtering steps included the removal of SNPs with a call rate <95%, MAF <0.01 and HWE P <1×10^−6^, as well as palindromic SNPs, duplicated SNPs, indels, and non-autosomal SNPs. After quality control, the number of variants totalled n = 252,170 CoreExome European, n = 217,508 CoreExome Pakistani, n = 615,108 CoreExome Bangladeshi, n = 471,297 GSA European, n = 406,938 GSA Pakistani, and n = 510,879 GSA Bangladeshi. Imputation of samples was performed using the Michigan imputation server (Minimac4)^25,26^. Phasing was performed using Eagle v2.4^35^. Imputation was performed for each ancestry group separately, all using the HRC v1.1 as the reference panel^27^. After imputation variants with poor estimated imputation accuracy (R^2^ <0.3) were removed.

#### Exeter Family Study of Childhood Health (EFSOCH)

The EFSOCH study is a prospective study of children born between 2000 and 2004, and their parents, from a geographically defined region of Exeter, UK. Participants were selected to comprise a homogeneous, European-ancestry cohort^36^. Pregnant women and their partners attended the study clinic when the women were 28 weeks pregnant, where anthropometric measures were collected. Additionally, a fasting blood sample was obtained from both parents at this visit and DNA was extracted, whilst offspring DNA was obtained from cord blood at birth^36^.

Anthropometric measures were taken on the children within 12 hours of delivery, with weight being recorded to the nearest 10 grams^36^. Maternal blood pressure measurements taken over the course of pregnancy were extracted and collated from clinical records retrospectively; therefore, like in the BiB study, multiple measures were available for each participant with numerous measures sometimes taken within the same gestational week and not always precisely at 28 weeks. We therefore employed the same method to select a blood pressure measure closest to 28 weeks gestation as we did for the BiB cohort.

Genotyping of 2,768 EFSOCH samples (n = 969 mothers, 937 fathers and 862 children) was performed using the Illumina Infinium HumanCoreExome-24 array (n = 551,839 SNPs/indels)^31^. QC procedures involved exclusion based on DNA sample call rate <98% (n = 50 individuals excluded), SNP call rate <95% (n = 13,151 SNPs excluded), minor allele frequency <1% (n = 257,289 further SNPs excluded), Hardy-Weinberg P <1×10^−6^ (n = 455 further SNPs excluded), sex mismatch between phenotypic sex and genotypically-derived sex (n = 13 individuals excluded), kinship errors after estimation using KING^21^ (n = 22 individuals excluded), and PC analysis outliers >4.56 SD from the European cluster mean after analysis to determine ancestry of the sample using flashPCA^37^ (n = 21 individuals excluded). Genotype imputation was subsequently performed using the Michigan imputation server^25,26^ with samples imputed to the Haplotype Reference Consortium HRC v1.1 reference panel^27^. If imputation quality score was >0.4 and MAF ≥1%, SNPs were retained for analysis. Following exclusions post-imputation, 7,674,771 SNPs were available for analysis in 2,662 individuals (939 mothers, 911 fathers and 812 children). As genotyping was performed in three batches, analyses were adjusted for batch.

#### Norwegian Mother, Father and Child Cohort Study (MoBa)

The Norwegian Mother, Father and Child Cohort Study (MoBa) is a population-based pregnancy cohort study conducted by the Norwegian Institute of Public Health^38^. Participants were recruited from all over Norway from 1999-2008^38^. The women consented to participation in 41% of the pregnancies^38^. The cohort includes approximately 114,500 children, 95,200 mothers and 75,200 fathers^38^. The current study is based on version 12 of the quality-assured data files.

Women were asked to complete three questionnaires, each at a different stage of pregnancy, covering data on past pregnancies, medical history, occupation/work factors, lifestyle factors, food frequency, and any changes in work situation or lifestyle habits^39^. Outcome data used in this study were collected via Questionnaire 3 (https://www.fhi.no/en/ch/studies/moba/for-forskere-artikler/questionnaires-from-moba/) sent out at 30 weeks of pregnancy, in which mothers were asked to report a blood pressure measure (the highest reading they had) only if they had been told they had high blood pressure currently or at some point during their pregnancy, with “high” defined as “over 140/90”. The date of this measurement and child’s gestational age at time of measure were not collected. As such, data were available for analysis as a continuous outcome, left-censored at 139 mmHg for SBP and 89 mmHg for DBP, or as a dichotomous outcome (reported high blood pressure or not). The study is linked with the Medical Birth Registry of Norway (MBRN)^40^, a national health registry containing information about all births in Norway^39^, where birth weight measures and information on maternal health prior to and during pregnancy were extracted from medical records.

Upon consent for participation at ultrasound exam, women were referred to the laboratory for blood sample collection^39^. After birth, a second blood sample was taken from the mother and umbilical cord blood collected from the child, from which DNA was extracted for genetic analysis^38^. In the current study, a subset of the MoBa cohort was used comprising maternal and fetal genotypes from 31,496 parent-offspring trios that were genotyped over 2012–2018^41,42^. Only singleton, live-birth pregnancies with complete birth registry data and at least the first MoBa questionnaire answered, as well as individuals alive at time of processing, were genotyped. Pregnancies were further excluded that used IVF, in cases of maternal diabetes, where the child’s APGAR score was 0 at both 1 and 5 minutes, and where recorded gestational duration was an extreme outlier. One random pregnancy was kept for each mother that had more than one pregnancy in the cohort^43^.

Genotyping was performed in nine batches on different Illumina arrays (HumanCoreExome-12 v1.1, HumanCoreExome-24 v1.0, Global Screening Array v1.0, InfiniumOmniExpress-24 v.2, HumanOmniExpress-24 v1.0). Genotypes were called using Illumina GenomeStudio software (v.2011.1 and v.2.0.3). Cluster positions were identified from samples with call rate ≥ 0.98 and GenCall score ≥ 0.15. Variants were excluded on the basis of GenomeStudio parameters of cluster separation < 0.4, 10 % GC-score < 0.3, AA T Dev > 0.025, call rate < 98%, or HWE P <1×10^−6^. Samples were excluded if they exhibited < 98% call rate, heterozygosity excess > 4 SD, genotyped sex mismatch, or were related to another sample in the cohort with estimated genome-wide identity by descent proportion (PI_HAT) > 0.1. Lastly, individuals of primarily non-European ancestry were excluded based on PCA with reference samples from the HapMap project, version 3^43^.

Pre-phasing was conducted locally using Shapeit v2.790^24^. Imputation was performed on the Sanger Imputation Server with positional Burrows-Wheeler transform and HRC version 1.1 reference panel^27^. All coordinates are provided in reference to the human genome build GRCh37^43^.

#### Observational associations between fetal sex and maternal blood pressure

We performed multivariable linear regression in each cohort to explore the association between fetal sex and maternal SBP and DBP measured at approximately 28 weeks gestation using term (≥37 weeks and <43 weeks), singleton pregnancies with live births in unrelated women according to two main models:

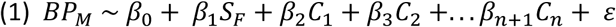

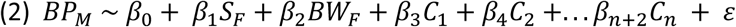

where *BP_M_* is maternal blood pressure (SBP or DBP) in pregnancy, *S_F_* is fetal sex, and *BW_F_* is a standardized variable for fetal birth weight obtained by calculation of a z-score i.e. subtracting the mean value (m) of birth weight, then dividing by the standard deviation (sd) e.g. x = (x – m)/sd, which has not been adjusted for sex or gestational age. *C* represents additional study specific covariates, and ε is the residual error. Models were adjusted for the covariates maternal age, parity, self-reported ancestry where cohorts included non-Europeans (HAPO and BiB), and in the cohorts where a BP measure was selected from a range of values surrounding 28 weeks (BiB and EFSOCH), gestational age at time of measurement. As not all individuals had genetic information, sample size was maximized by using all available phenotypic data, therefore, self-reported ancestry was used here to adjust for ancestral population differences impacting the variance in our BP outcome rather than genetic PCs. We do not have reason to believe that any effect of fetal sex on maternal BP would differ in size between ancestry groups, therefore controlling for self-reported ancestry in each cohort was sufficient, with no need to model each ancestry separately. For each cohort, the sex effect estimates from the two models were then informally compared to get an indication of whether any fetal sex effects observed were likely being driven by a difference in birth weight rather than an independent mechanism.

SBP measures below 80 mmHg and DBP measures below 50 mmHg were classified as outliers and excluded from each cohort. For each cohort, we used its sample size to estimate the number of standard deviations from the mean within which 95% of birth weight observations are expected to fall given the number of individuals in each cohort (EFSOCH = 3, HAPO = 4, ALSPAC = 4, BiB = 4 SDs, respectively). Birth weight observations outside of this range were classified as outliers and excluded prior to analysis (observations excluded: EFSOCH = 1, HAPO = 474, ALSPAC = 156, BiB = 581). For the MoBa sample, outliers were determined based on precalculated birth weight z-scores for gestational age, removing those with magnitude above five standard deviation units. All analyses were first performed within each individual study population and then meta-analysed using a fixed-effects model to obtain the fetal sex effect across all cohorts, comprising a total sample size of up to 109,842 mother-child pairs (HAPO n = 12,052; ALSPAC n = 10,374; BiB n = 6,486; EFSOCH n = 784; MoBa n = 80,146). For analysis of the censored MoBa data, the survreg function in R package ‘survival’^44^ was used to fit a left-censored regression model with a Gaussian distribution.

Analyses were repeated using logistic regression to also evaluate the data in closer alignment with the way data was collected for the MoBa study, which comprised our largest sample, adhering to the same inclusion and exclusion criteria as above. This dichotomous ‘high BP’ outcome variable was generated according to clinical criteria for hypertension^45^ and in accordance with the MoBa questionnaire, partitioning measures based on whether SBP or DBP were ≥140 mmHg and 90 mmHg, respectively. All analyses were again performed first within each cohort and then fixed-effect meta-analysed to obtain the fetal sex effect estimate across all studies. Association analyses were performed in MoBa using R version 4.1.2 and in the remaining cohorts using Stata version 16.1^46^, and all meta-analysed using the rma function in R package ‘metafor’^47^, R version 4.1.2^48^.

### Fetal genetic scores for birth weight

Unweighted fetal genetic scores were constructed for birth weight and tested for an association with maternal SBP, and DBP measured in term, singleton pregnancies from unrelated women at approximately 28 weeks gestation. The genetic data was obtained using mother-child pairs of European ancestry from the HAPO (n = 474), ALSPAC (n = 4,993), EFSOCH (n = 539), BiB (n = 1,720), and MoBa (n = 22,949) cohorts, and individuals of South Asian ancestry (Pakistani n = 1,501 and Bangladeshi n = 56) within the BiB cohort.

The genetic scores were calculated using 186 autosomal own-birth-weight-associated lead SNPs from the marginal analysis from Warrington et al. 2019 genome-wide association study (GWAS) for birth weight^49^, see S1 Table. The genetic score (GS) was calculated according to the following equation,

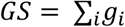

where *g_i_* is the genotype (dosage of birth-weight-raising-alleles 0, 1, or 2) at SNP *i* summed across all birth-weight-associated SNPs for each individual in the sample. The calculated genetic score was then standardized, and the maternal outcomes regressed on the standardised fetal genetic score in independent linear regression models using Stata version 16.1^46^. The South Asian ancestries within the BiB sample were modelled separately, and effects were meta-analyzed across all cohorts with available measures for each outcome using a random effects model to account for any heterogeneity in effect sizes between sample ancestries, performed with the rma function in R package ‘metafor’^47^, R version 4.1.2^48^. All genetic score association models with maternal BP were adjusted for a standardized unweighted maternal genetic score for birth weight constructed in the same manner as above to control for the correlated maternal genotype^50^, and where possible, the first 5 maternal ancestry principal components to adjust for potential population structure affecting the outcome variable (except in MoBa, where the first 3 fetal PCs were used which are strongly correlated with maternal PCs), and additionally for genotyping batch effects in the EFSOCH and MoBa studies.

## Results

The characteristics of each cohort are described in **Table 1**. Participants within the cohorts were largely consistent in many of the characteristics including mean offspring birth weight, distribution of sex amongst offspring, gestational age at birth, maternal height, and maternal age, with mothers in BiB being on average slightly younger. Mothers in the HAPO and BiB studies had higher mean BMIs as the measurements for these women were taken at the time of an OGTT at approximately 28 weeks gestation, whereas pre-pregnancy measurements were obtained for the women in the other cohorts. Women in HAPO also had the lowest mean measures of SBP and were predominantly primiparous relative to the women of the other cohorts who were fairly evenly split between primiparous and multiparous. Cohorts were largely of White ethnicity aside from HAPO and BiB where a more ancestrally diverse sample was available.

**Table 1.**
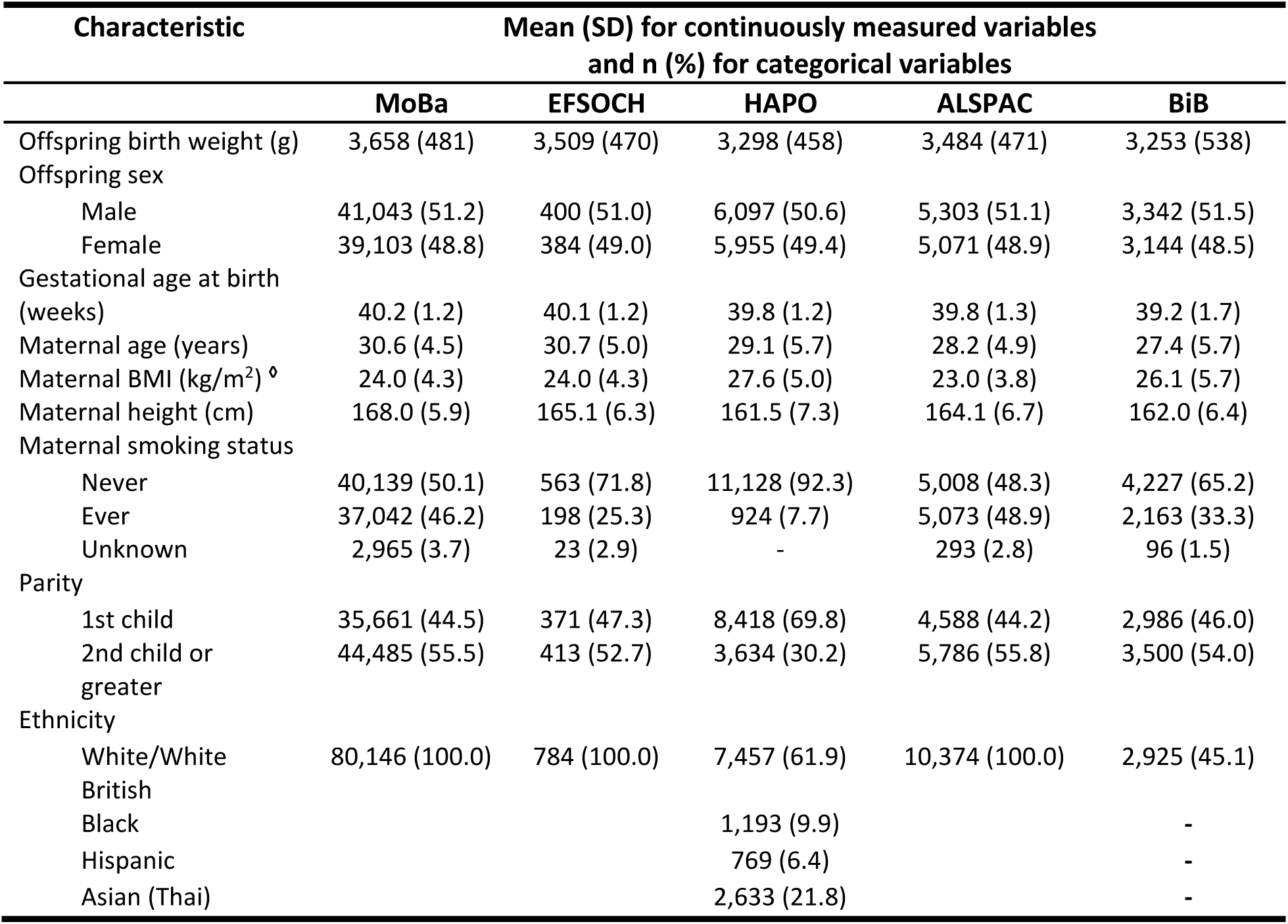
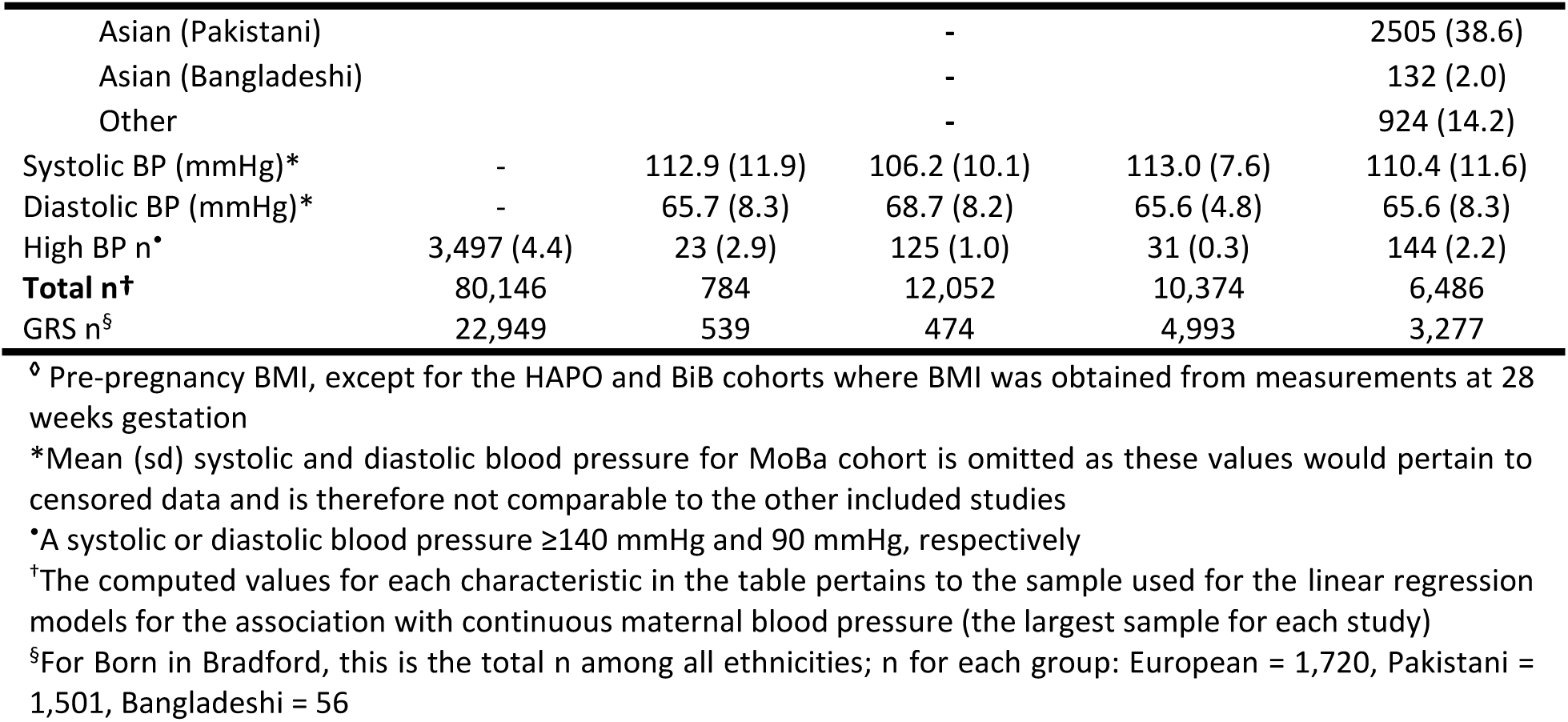
Descriptive statistics of cohort participants’ traits and characteristics.

### Fetal sex and maternal blood pressure – continuous

After meta-analysis of individual fetal sex effects across cohorts, both maternal SBP and DBP were higher in pregnancy when carrying a male fetus compared to a female fetus (a mean difference of 0.35 mmHg [95%CI: 0.15-0.55] and 0.35 mmHg [95%CI: 0.21-0.49], for SBP and DBP, respectively]; **Figure 1a and b**). When birth weight was added to each model, evidence for an independent effect of fetal sex remained but attenuated slightly for both SBP and DBP (0.22 mmHg [95%CI: 0.02-0.42] and 0.31 mmHg [95%CI: 0.17-0.45], respectively; **Figure 2**).

**Figure 1.**
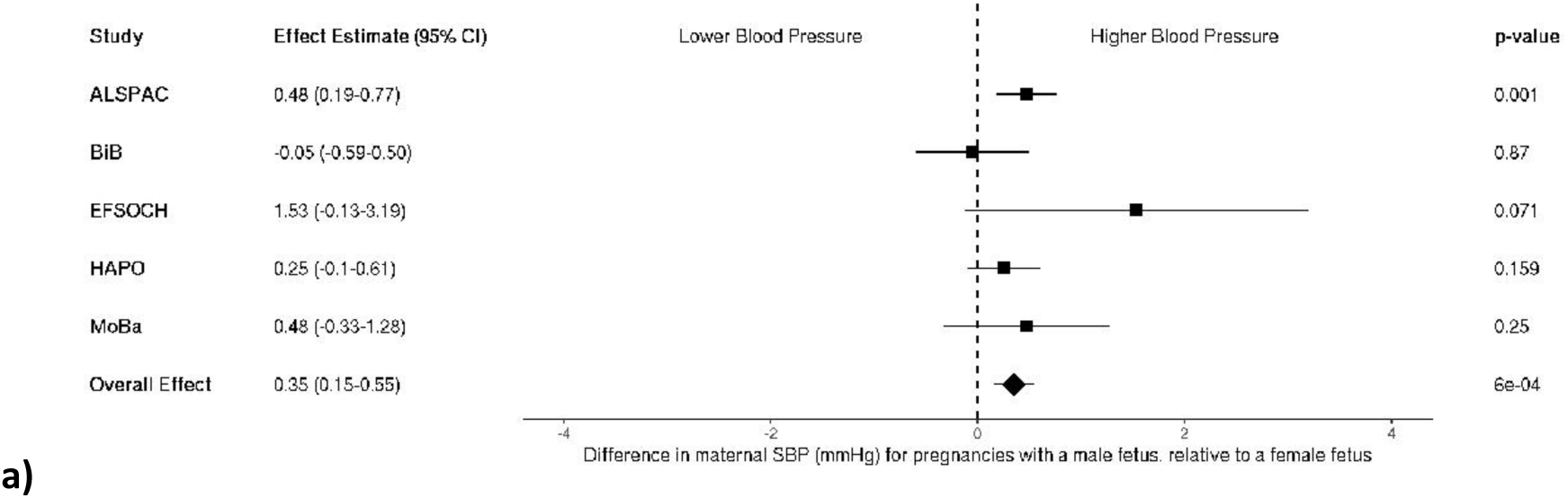
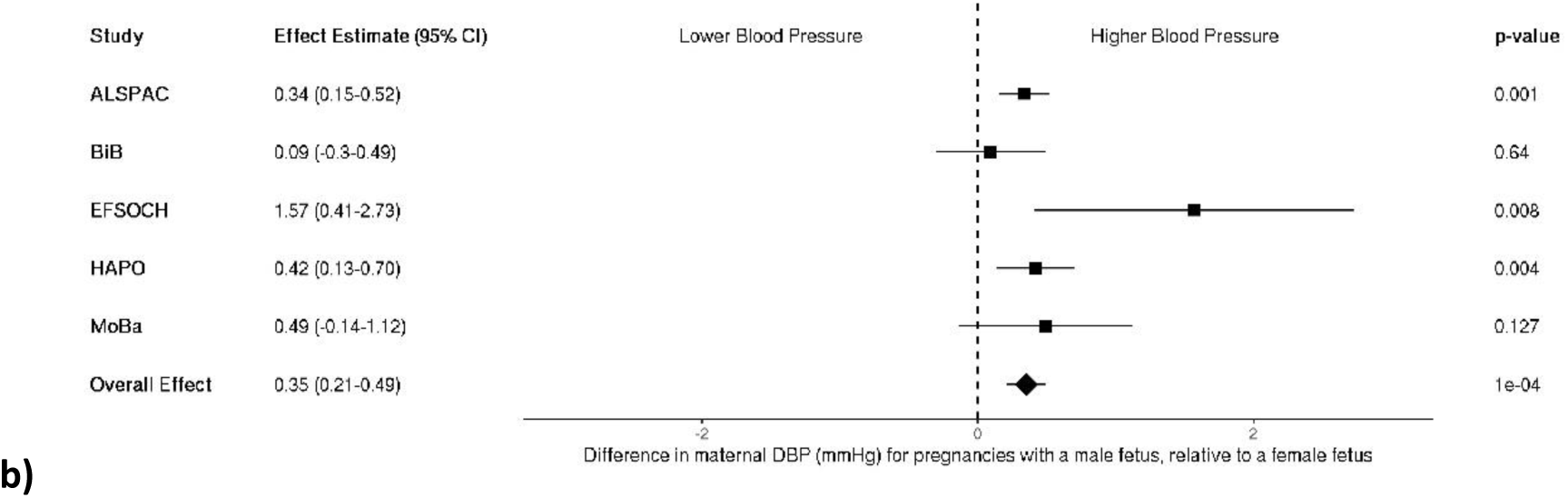
**a and b**. Meta-analysis of the fetal sex association with maternal a) systolic (n = 31,928) and b) diastolic blood pressure (n = 31,199). Each estimate is the difference in maternal blood pressure (mmHg) for pregnancies with a male fetus relative to a female fetus (95% CI). ALSPAC, Avon Longitudinal Study of Parents and Children; BiB, Born in Bradford study; EFSOCH, Exeter Family Study of Childhood Health; HAPO, Hyperglycemia and Adverse Pregnancy Outcome study; MoBa, Norwegian Mother, Father and Child Cohort Study; SBP, systolic blood pressure; DBP, diastolic blood pressure.

**Figure 2.**
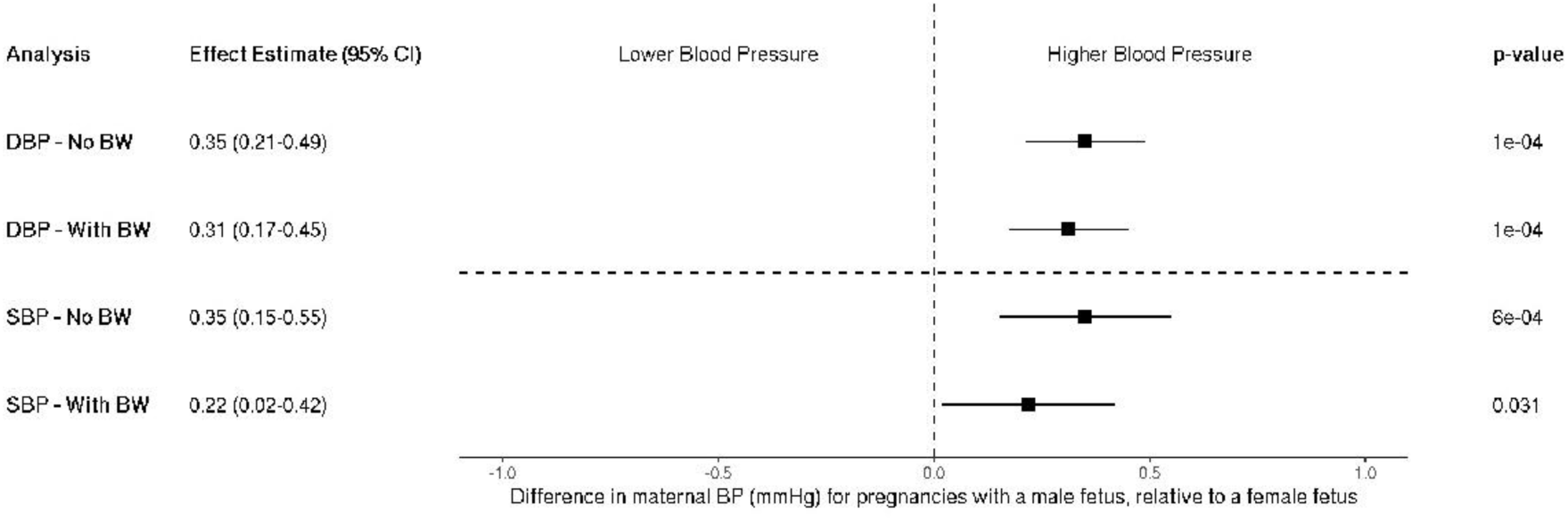
Meta-analyzed fetal sex associations with maternal systolic (n = 31,928) and diastolic blood pressure (n = 31,199), with and without adjustment for child’s birth weight. Each estimate is the difference in maternal blood pressure (mmHg) for pregnancies with a male fetus relative to a female fetus (95% CI). SBP, systolic blood pressure; DBP, diastolic blood pressure; BW, birth weight.

### Fetal sex and maternal blood pressure – logistic (high BP)

When dichotomizing maternal blood pressure into high and low categories, logistic regression analyses showed a positive effect estimate for odds of experiencing high maternal BP given pregnancy with a male fetus, but the confidence intervals were wide and crossed the null (OR 1.05 [95%CI: 0.98-1.12]; Figure 3].

**Figure 3.**
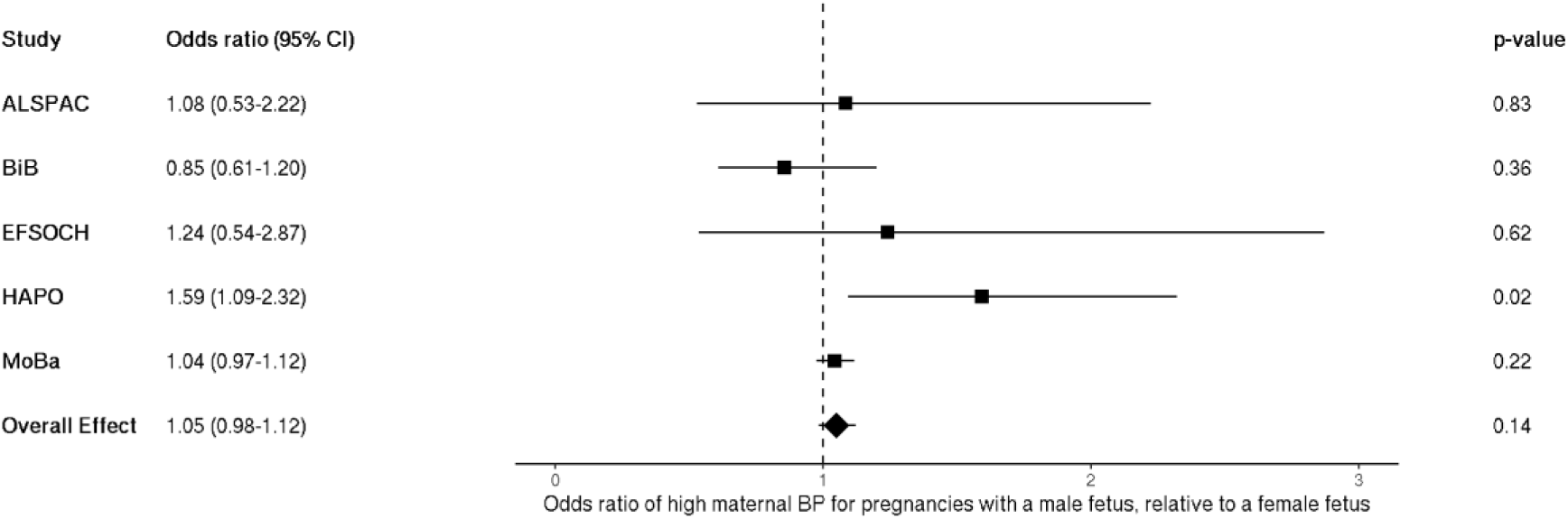
Meta-analysis of the fetal sex association with high maternal blood pressure, n = 109,842. Each estimate is the odds ratio of high maternal blood pressure for pregnancies with a male fetus relative to a female fetus (95% CI). ALSPAC, Avon Longitudinal Study of Parents and Children; BiB, Born in Bradford study; EFSOCH, Exeter Family Study of Childhood Health; HAPO, Hyperglycemia and Adverse Pregnancy Outcome study; MoBa, Norwegian Mother, Father and Child Cohort Study; BP, blood pressure.

### Birth weight genetic score and maternal blood pressure

When testing for an association between a fetal birth weight genetic score and maternal BP, we found no strong evidence for an association with SBP (0.09 mmHg per 1 SD change in fetal BW score [95%CI: −0.11-0.29]), or DBP (0.09 mmHg per 1 SD change in fetal BW score [95% CI: −0.05-0.22]; Figure 4a and b). When looking at maternal BP classified as high vs. low, there was no strong evidence of an association of high maternal BP in pregnancy with a fetal birth weight genetic score (OR 1.01 per 1 SD change in fetal BW score [95%CI: 0.94-1.08]; Figure 5).

**Figure 4.**
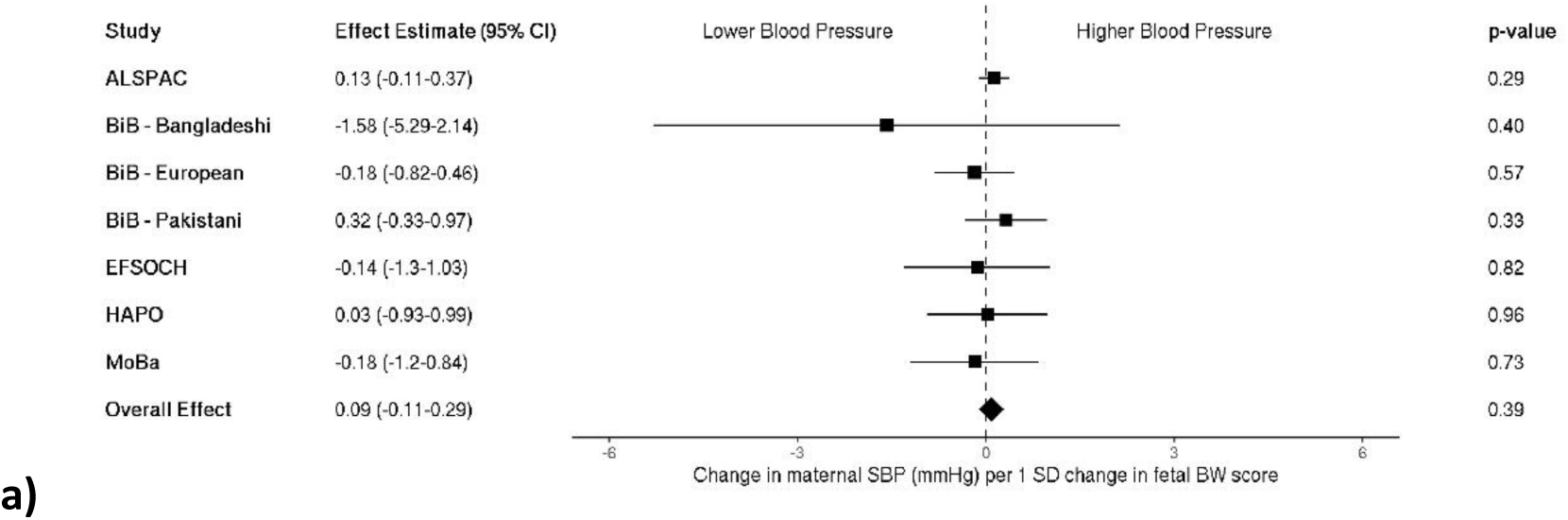
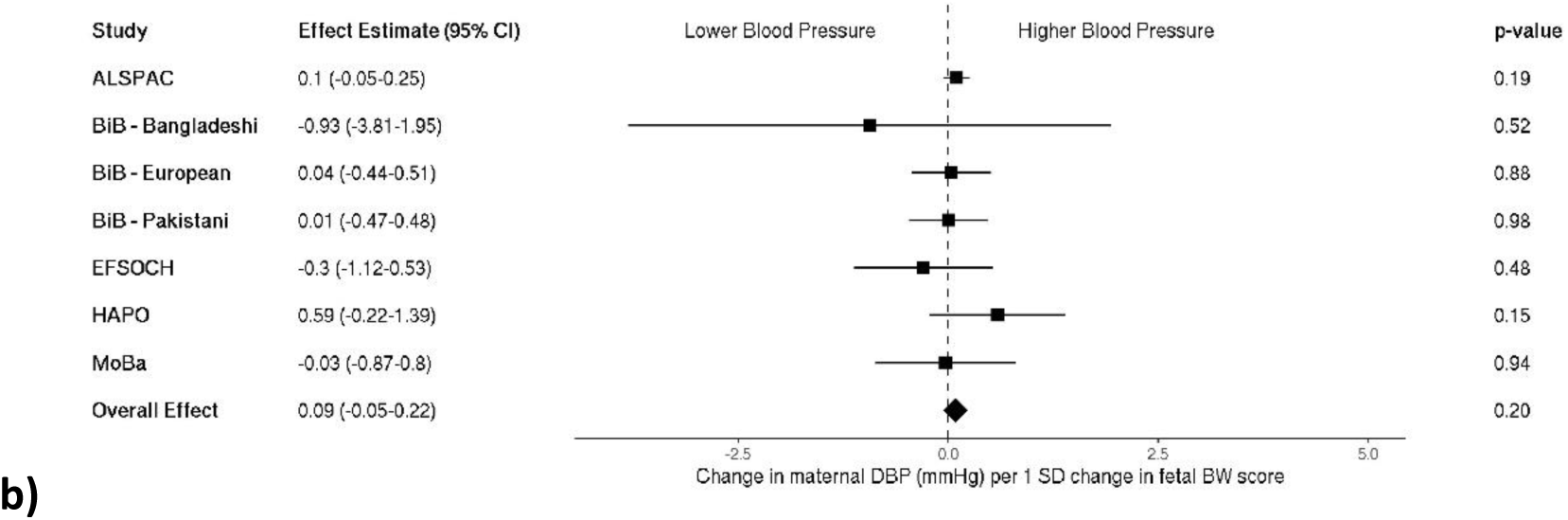
**a and b**. Meta-analysis of the fetal birth weight genetic score association with maternal a) systolic (n = 9,892) and b) diastolic blood pressure (n = 9,682). Each estimate is the change in maternal blood pressure (mmHg) per 1 standard deviation change in fetal birth weight score (95% CI). ALSPAC, Avon Longitudinal Study of Parents and Children; BiB, Born in Bradford study; EFSOCH, Exeter Family Study of Childhood Health; HAPO, Hyperglycemia and Adverse Pregnancy Outcome study; MoBa, Norwegian Mother, Father and Child Cohort Study; SBP, systolic blood pressure; DBP, diastolic blood pressure; SD, standard deviation; BW, birth weight.

**Figure 5.**
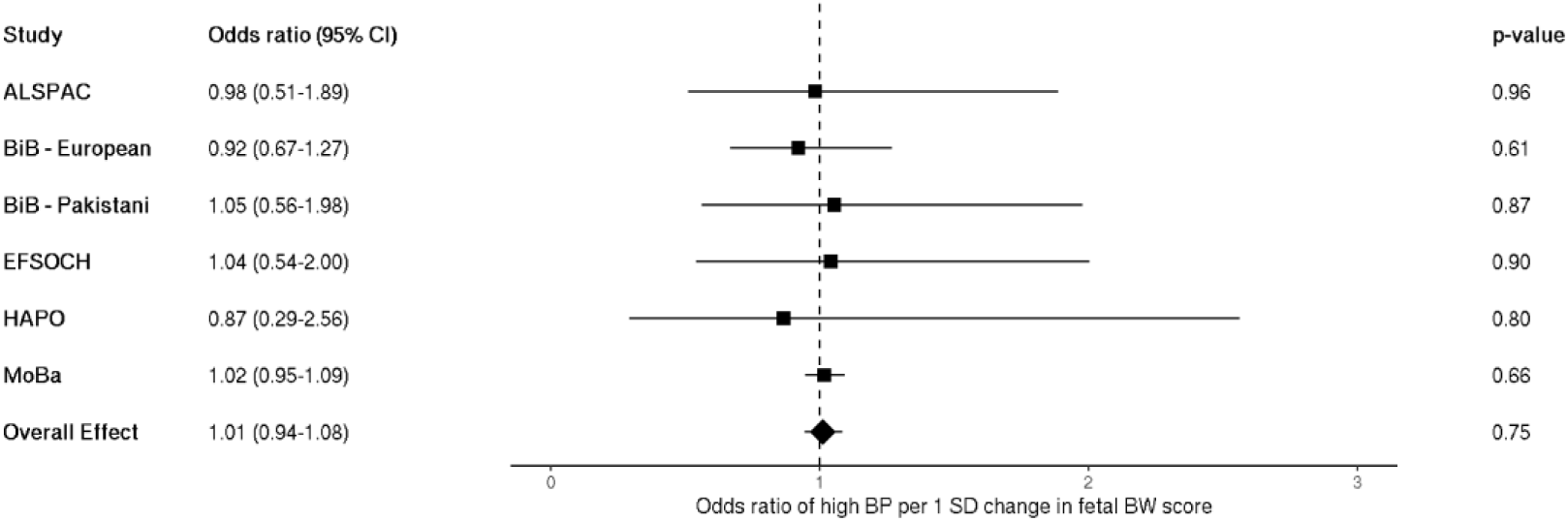
Meta-analysis of the fetal birth weight genetic score association with high maternal blood pressure; n = 32,232. Each estimate is the odds ratio for high blood pressure per 1 standard deviation change in fetal birth weight score (95% CI). ALSPAC, Avon Longitudinal Study of Parents and Children; BiB, Born in Bradford study; EFSOCH, Exeter Family Study of Childhood Health; HAPO, Hyperglycemia and Adverse Pregnancy Outcome study; MoBa, Norwegian Mother, Father and Child Cohort Study; BP, blood pressure; SD, standard deviation; BW, birth weight.

## Discussion

Overall, we found strong evidence to support an effect of male fetal sex on higher maternal BP in pregnancy. When adjusting for birth weight, and thus the inherent difference in size between male and female babies, the effect of fetal sex attenuated slightly, although more so for maternal SBP than DBP. This may suggest that the small increases in maternal BP associated with male fetal sex are not primarily driven by males being heavier and that a mechanism altering this aspect of maternal physiology during pregnancy through fetal sex exists independent of birth weight. In a secondary analysis, we assessed this relationship using a dichotomous outcome where women were assigned to groups of ‘high BP’ or not. Compared to the overall sample, there were few cases of ‘high BP’ (n = 3,820) and thus low statistical power despite the larger overall sample size, though the results were directionally consistent with our continuous outcome findings. The estimates obtained in this study give an indication of any increased clinical risk associated with having a male baby (which is small).

Given the attenuation in the fetal sex effect when adjusting for birth weight, we sought to test for an association between a fetal genetic score for birth weight and maternal BP to better determine whether birth weight could be mediating some of the effect of sex. In our study, we did not find evidence for an association between a fetal birth weight genetic score and maternal BP in pregnancy. One study published by Chen et al. investigated this relationship previously and found that fetal alleles that increase birth weight are associated with a higher BP in the mother and a shorter gestational duration^16^. Though we did not find the same, the effect estimates were directionally consistent with Chen et al. (0.09 mmHg higher SBP per 1 SD change in fetal BW score, *P* = 0.39 vs. 1.4 mmHg increase in SBP per 1-SD change in fetal growth rate, *P* = 0.023, respectively). There are some key differences between our methods of analysis and those used in their study, notably in how the genetic scores were composed. We included 186 autosomal SNPs associated with birth weight in an unweighted score, while Chen et al. constructed haplotype genetic scores independently for maternally and paternally inherited alleles using only 86 SNPs associated with birth weight with confirmed fetal effects, weighted by SEM-adjusted fetal effect sizes from the Warrington et al. GWAS. The use of maternal and paternal transmitted and non-transmitted haplotypes by Chen et al. controlled for the correlation between maternal and fetal genotype in their analysis, because under random mating the paternally transmitted fetal haplotype for birth weight should not be correlated with maternal genotype. We used the full set of birth weight loci to maximise power. With many variants included, rather than adjust for maternal genotype at each fetal genetic score locus separately in our models, we took an unweighted score approach, generating a fetal unweighted genetic score for birth weight and adjusting for maternal genotype at those same loci by including a maternal unweighted genetic score for birth weight in the models^50^.

Further comparisons between our study and that of Chen et al. concern the cohorts shared between our studies, ALSPAC and HAPO. Though we share the inclusion of the HAPO cohort, we have used non-overlapping samples, where our sample of individuals were genotyped locally in Exeter, UK, while their individuals came from a central study sample that did not include the Exeter sample at the time of their analysis. Mother-child pairs included from ALSPAC in our study (SBP/DBP n = 4,993) almost completely overlapped with those included in Chen et al.’s study (SBP n = 4,923, DBP n = 4,943). We did however use slightly different BP measurements in ALSPAC. Our study used maternal BP predicted at 28 weeks gestation for ALSPAC women allowing us to have a consistent time point with HAPO and our other included cohorts who were similarly measured at ∼28 weeks, while Chen et al. used the average of all BPs in ALSPAC measured between 30 to 36 weeks gestation^16^. Overall, despite non-uniform methods employed between the two studies, we would still expect both methods of analysis to obtain a similar result if there was a strong effect of child’s birth weight on maternal BP. Replication with greater power in the future, as well as the use of alternative metrics capturing fetal growth, may elucidate this further.

Our finding regarding fetal sex is consistent with our hypothesis that male sex is associated with higher maternal BP. However, our results cannot exclude the possibility of a reverse-causal association, where maternal BP before pregnancy somehow influences the sex of the baby. Such findings were reported by Retnakaran et al.^51^, who conducted a prospective preconception cohort study of approximately 1,400 newly married Chinese women who had cardiometabolic measures assessed prior to an eventual first pregnancy. They found that SBP before pregnancy was higher in women who delivered a boy than in those who delivered a girl, even after adjustment for covariates (106.0 vs. 103.3 mmHg, *P* = 0.0015). However, when following a proportion of the women with BP measured throughout their pregnancy, this difference in BP between mothers of boys and girls was not observed in any trimester of pregnancy. While this observational association raises the possibility that maternal pre-pregnancy BP could affect offspring sex, the association was not observed during pregnancy. Associations such as these highlight the need for further studies to support these findings and definitively determine the direction of influence between mother and fetus for this relationship.

A prominent example of fetal drive on maternal BP in pregnancy, and what helped form the fetal drive hypothesis, is that mothers of children with Beckwith-Wiedemann syndrome who give birth to macrosomic babies experience higher rates of gestational hypertension^11^. Further evidence that originally spurred the hypothesis came from the suggestion of a fetal genetic effect on maternal BP in animal models of mice exhibiting loss of the imprinted p57*^kip^*^2^ gene, an inhibitor of a variety of cyclin/cyclin-dependent kinase (CDK) complexes which regulate the cell cycle of placental trophoblasts and other cell types^52^. In the study, half of the unborn offspring were p57*^kip^*^2^ deficient and exhibited enlarged placentas and growth restriction. Their functionally wild type mothers exhibited pre-eclampsia-like symptoms of pregnancy-induced hypertension and proteinuria in late pregnancy which went away after birth.

Additional evidence has been found in the literature for a relationship between fetal factors and hypertensive disorders in human pregnancy. A systematic review and meta-analysis by Broere-Brown et al. found that fetal sex was associated with pre-eclampsia, a disorder characterized by placental insufficiency, in a gestational age dependent manner, where the occurrence of preterm pre-eclampsia was greater when carrying a female fetus and the association reversed when carrying a male fetus with greater occurrence of term and post-term pre-eclampsia^6^. The authors theorized that this trend suggests that males that are not viable due to impaired placentation will be lost early while those that make it to term, though with late-onset pre-eclampsia, represent a comparatively heathy subset. This idea is supported by Orzack et al.’s human sex ratio study where an excess male mortality was observed within approximately the first week following conception, followed by greater female-biased mortality until approximately 20 weeks gestation, and then excess male mortality towards the end of pregnancy^53^. Additionally, in a recent GWAS of placental weight (PW), causal methods indicated that a fetal genetic predisposition to a higher PW increases the risk of pre-eclampsia^54^. While an association with sex was not investigated in that study, the findings serve as an additional example of fetal genetic influence on maternal physiology and BP-related outcomes.

The fetal sex association results of our study contribute an example in healthy human pregnancies of fetal effects on the mother. We do, however, acknowledge some important limitations. We know that male babies grow larger and faster *in utero* than female babies^9,10^, and that birth weight alone, as we have assessed here, does not entirely capture fetal growth and the full extent of growth differences between males and females, nor does it fully capture their effects on the mother. Therefore, despite the lack of association found between genetically determined birth weight and maternal BP, some of the residual differences between males and females in our study may still be due to growth differences that should be investigated further. It should be noted that our fetal genetic score for birth weight likely accounts for only a small proportion of variation in birth weight (including, presumably, genetically determined variation in birth weight), which may also contribute to the absence of an association. A further limitation in our study is that birth weight is likely to be a collider variable along the path between our exposure (fetal sex) and outcome of interest (maternal blood pressure), making interpretation uncertain as adjusting for it in our analysis could lead to the opening of a pathway of association from some unmeasured variable that affects both birth weight and maternal BP to our exposure of interest, fetal sex i.e. collider bias^55^. However, given the consistency of our estimates when adjusting for birth weight or not, it is unlikely that this is impacting our results.

Additionally important to note is that despite the overall sample sizes achieved, we are likely underpowered in the analysis of maternal hypertension as there was a bias toward healthy women in the included studies with comparatively few women that met the threshold for high BP. Within the high BP group, we also must acknowledge the general pitfalls of self-reporting coming from the MoBa study (i.e. the largest contributing sample in our study), where there are varying time points for when BP would have been measured during each pregnancy compared to if collection took place at a specified clinic visit, as well as variation in quality of response e.g. whether the measure given adhered to the questionnaire definition of high BP or not and how this contributed to how women were grouped for analysis. For the continuous maternal BP analyses, the data were censored at the defined threshold for high BP, however, for the dichotomous BP outcome, women were sorted into the ‘high BP’ group so long as they answered ‘yes’ to the initial binary question of whether they had, or had had, high BP during their pregnancy. Thus the ‘high BP’ group contained some women who answered ‘yes’ but then provided a measure below the threshold.

Lastly, our study was performed in a largely White European ancestry sample, where non-Europeans comprised only 7.4% of the total study sample size (n = 8,156 / 109,842). Though our results did not indicate substantial differences between ancestral/ethnic groups across our analyses, larger samples of non-European individuals may provide greater power to detect differences in strength of association and more precise estimates in diverse populations in the future.

In conclusion, we found that carrying a male fetus is associated with a higher maternal BP in pregnancy relative to carrying a female fetus, and through both observational and genetic analyses investigating a role for birth weight, our results suggest that larger fetal size at birth does not account for a substantial part of this association. The residual association may be attributable to faster male growth that is not captured by birth weight or may be partially driven by an independent mechanism related to fetal sex. Our findings do not point to a difference in maternal BP that would warrant significant changes to routine monitoring in clinical practice, but they do suggest that male sex may contribute as a risk factor for BP-related complications. We add here to the body of literature supporting the hypothesis that sexual dimorphism exists within the spectrum of adverse pregnancy outcomes and that male fetuses may present greater cardiometabolic challenges to their mothers compared to female counterparts. Future research would benefit from further observational studies looking at fetal sex and its association with other maternal physiological outcomes within the normal range, not only in the disease state, and especially in more ancestrally diverse populations.

## Data Availability

Summary statistics from EFSOCH are available on request. Researchers interested in accessing the data are expected to send a reasonable request by sending an email to the Exeter Clinical Research Facility at.
For access to the HAPO data used in this study, please contact Dr Rachel Freathy and Prof. William Lowe Jr. The website describing the study and other data available is https://www.ncbi.nlm.nih.gov/projects/gap/cgi-bin/study.cgi?study_id=phs000096.v4.p1
The ALSPAC data management plan describes in detail the policy regarding data sharing, which is through a system of managed open access. The data used in this study are linked to ALSPAC project number B3392. To request access to the data included in this paper and all other existing ALSPAC data: (i) Please read the ALSPAC access policy, which describes the process of accessing the data and samples in detail and outlines the costs associated with doing so, (ii) you may also find it useful to browse the fully searchable ALSPAC research proposals database, which lists all research projects that have been approved since April 2011, and (iii) please submit your research proposal for consideration by the ALSPAC Executive Committee. You will receive a response within 10 working days to advise you whether your proposal has been approved. If you have any questions about accessing data, please. Please note that the study website contains details of all the data that is available through a fully searchable data dictionary and variable search tool: http://www.bristol.ac.uk/alspac/researchers/our-data/.
Scientists are encouraged and able to use BiB data. Data requests are made to the BiB executive using the form available from the study website http://www.borninbradford.nhs.uk (please click on Science and Research to access the form). Guidance for researchers and collaborators, the study protocol and the data collection schedule are all available via the website. All requests are carefully considered and accepted where possible.
Data from the Norwegian Mother, Father and Child Cohort Study and the Medical Birth Registry of Norway used in this study are managed by the Norwegian Institute of Public Health and are available to researchers through an application via https://helsedata.no. The consent given by the participants does not open for storage of data on an individual level in repositories or journals. Access to data sets requires approval from a Regional Committee for Medical and Health Research Ethics in Norway and an agreement with MoBa.

http://www.bristol.ac.uk/alspac/researchers/our-data/

http://www.borninbradford.nhs.uk/

https://helsedata.no/

## Supporting information

**S1 Table. 186 autosomal own-birth-weight-associated lead SNPs, oriented to the increaser allele.** From the marginal analysis of Warrington et al. 2019 genome-wide association study for birth weight.

## Data availability

Summary statistics from EFSOCH are available on request. Researchers interested in accessing the data are expected to send a reasonable request by sending an email to the Exeter Clinical Research Facility at.

For access to the HAPO data used in this study, please contact Dr Rachel Freathy and Prof. William Lowe Jr. The website describing the study and other data available is https://www.ncbi.nlm.nih.gov/projects/gap/cgi-bin/study.cgi?study_id=phs000096.v4.p1

The ALSPAC data management plan describes in detail the policy regarding data sharing, which is through a system of managed open access. The data used in this study are linked to ALSPAC project number B3392. To request access to the data included in this paper and all other existing ALSPAC data: (i) Please read the ALSPAC access policy, which describes the process of accessing the data and samples in detail and outlines the costs associated with doing so, (ii) you may also find it useful to browse the fully searchable ALSPAC research proposals database, which lists all research projects that have been approved since April 2011, and (iii) please submit your research proposal for consideration by the ALSPAC Executive Committee. You will receive a response within 10 working days to advise you whether your proposal has been approved. If you have any questions about accessing data, please. Please note that the study website contains details of all the data that is available through a fully searchable data dictionary and variable search tool: http://www.bristol.ac.uk/alspac/researchers/our-data/.

Scientists are encouraged and able to use BiB data. Data requests are made to the BiB executive using the form available from the study website http://www.borninbradford.nhs.uk (please click on ‘Science and Research’ to access the form). Guidance for researchers and collaborators, the study protocol and the data collection schedule are all available via the website. All requests are carefully considered and accepted where possible.

Data from the Norwegian Mother, Father and Child Cohort Study and the Medical Birth Registry of Norway used in this study are managed by the Norwegian Institute of Public Health and are available to researchers through an application via https://helsedata.no. The consent given by the participants does not open for storage of data on an individual level in repositories or journals. Access to data sets requires approval from a Regional Committee for Medical and Health Research Ethics in Norway and an agreement with MoBa.

## Ethics approval

Ethical approval for the Exeter Family Study of Childhood Health was given by the North and East Devon (UK) Local Research Ethics Committee (approval number 1104), and informed consent was obtained from the parents of the newborns.

Ethical approval for the study was obtained from the ALSPAC Ethics and Law Committee and the Local Research Ethics Committees. Informed consent for the use of data collected via questionnaires and clinics was obtained from participants following the recommendations of the ALSPAC Ethics and Law Committee at the time. Consent for biological samples has been collected in accordance with the Human Tissue Act (2004). Study participants have the right to withdraw their consent for elements of the study or from the study entirely. Full details of the ALSPAC consent procedures are available on the study website (http://www.bristol.ac.uk/alspac/researchers/research-ethics/).

Ethics approval was obtained for the main platform study and all of the individual sub-studies from the Bradford Research Ethics Committee.

The establishment of MoBa and initial data collection was based on a licence from the Norwegian Data Protection Agency and approval from The Regional Committees for Medical and Health Research Ethics. The MoBa cohort is currently regulated by the Norwegian Health Registry Act. The administrative board of MoBa led by the Norwegian Institute of Public Health approved the study protocol and the current study was approved by the Regional Committees for Medical and Health Research Ethics (no. 2012/67).

## Author contributions

C.S.D., R.N.B., N.M.W., K.A.P., A.T.H., D.M.E., and R.M.F. contributed to the conceptualization and/or design of the study. C.S.D. performed the analyses in this study with special contribution by J.J. who performed the analysis in the MoBa cohort. Data interpretation and statistical analysis was aided by R.N.B., J.J., N.M.W., D.M.E., and R.M.F. S.J., P.R.N. and B.J., and W.L.L. were involved in data curation for the MoBa and HAPO studies, respectively. C.S.D. drafted the original manuscript including all tables and figures. All authors reviewed and edited versions of the manuscript. All authors read and approved the final manuscript. This work was jointly directed by R.N.B. and R.M.F.

## Acknowledgements

This study represents independent research supported by the National Institute of Health Research (NIHR) Exeter Clinical Research Facility. The views expressed are those of the author(s) and not necessarily those of the NHS, the NIHR or the Department of Health and Social care.

The Exeter Family Study of Childhood Health (EFSOCH) was supported by South West NHS Research and Development, Exeter NHS Research and Development, the Darlington Trust and the Peninsula NIHR Clinical Research Facility at the University of Exeter. The opinions given in this paper do not necessarily represent those of NIHR, the NHS or the Department of Health. We would like to acknowledge Andrew Hattersley as the principal investigator, and Bea Knight for her contribution to data collection, of the EFSOCH study.

We are extremely grateful to all the families who took part in this study, the midwives for their help in recruiting them, and the whole ALSPAC team, which includes interviewers, computer and laboratory technicians, clerical workers, research scientists, volunteers, managers, receptionists and nurses.

Born in Bradford is only possible because of the enthusiasm and commitment of the children and parents in BiB. We are grateful to all the participants, health professionals, schools and researchers who have made Born in Bradford happen.

The Norwegian Mother, Father and Child Cohort Study is supported by the Norwegian Ministry of Health and Care Services and the Ministry of Education and Research. We are grateful to all the participating families in Norway who take part in this on-going cohort study.

