## Supplemental Table S1 for "The influence of fetal sex on maternal blood pressure in pregnancy"

**S1 Table. 186 autosomal own-birth-weight-associated lead SNPs, oriented to the increaser allele.** From the marginal analysis of Warrington et al. 2019 genome-wide association study for birth weight.

| SNP | Chrom:position (hg19) | Trait-raising Allele | Trait-lowering Allele | Beta (SEM[a] adjusted fetal effects) |
|---|---|---|---|---|
| rs17367504 | chr1:11862778 | A | G | 0.005 |
| rs12401656 | chr1:43456767 | G | A | 0.029 |
| rs80278614 | chr1:119412317 | A | G | 0.052 |
| rs905938 | chr1:154991389 | C | T | 0.023 |
| rs670523 | chr1:155878732 | G | A | 0.016 |
| rs72480273 | chr1:161644871 | C | A | 0.022 |
| rs10913200 | chr1:176521655 | G | A | 0.038 |
| rs61830764 | chr1:212289976 | A | G | 0.018 |
| rs3806315 | chr1:214724668 | A | G | 0.016 |
| rs708122 | chr1:228216997 | C | A | 0.015 |
| rs10495563 | chr2:9662210 | A | G | 0.016 |
| rs2551347 | chr2:23912401 | T | C | 0.029 |
| rs1179494 | chr2:36809496 | G | C | 0.002 |
| rs754868 | chr2:43185532 | G | A | 0.019 |
| rs4952673 | chr2:43423870 | G | A | 0.004 |
| rs17034876 | chr2:46484310 | T | C | 0.039 |
| rs4953353 | chr2:46567276 | G | T | 0.019 |
| rs186606513 | chr2:97482001 | G | A | 0.047 |
| rs56188432 | chr2:158406865 | G | A | 0.25 |
| rs560887 | chr2:169763148 | T | C | 0.025 |
| rs2280235 | chr2:191843830 | G | A | 0.014 |
| rs10181515 | chr2:227019461 | T | C | 0.021 |
| rs9855896 | chr3:14287150 | A | G | 0.014 |
| rs2168443 | chr3:46947087 | T | A | 0.01 |
| rs11708067 | chr3:123065778 | G | A | 0.056 |
| rs9851257 | chr3:123125711 | T | A | 0.005 |
| rs6440006 | chr3:141142691 | G | A | 0.001 |
| rs2306700 | chr3:142123841 | T | C | 0.022 |
| rs10935733 | chr3:148622968 | T | C | 0.021 |
| rs4679760 | chr3:155855418 | C | G | 0.009 |
| rs1482852 | chr3:156798294 | A | G | 0.054 |
| rs11711420 | chr3:183349010 | T | G | 0.022 |
| rs4144829 | chr4:17903654 | C | T | 0.032 |
| rs6533183 | chr4:106133184 | C | T | 0.008 |
| rs116807401 | chr4:135121721 | C | T | 0.088 |
| rs6845999 | chr4:145565826 | T | C | 0.017 |
| rs4579095 | chr4:174726635 | A | G | 0.007 |
| rs1818782 | chr5:39424628 | C | A | 0.015 |
| rs351930 | chr5:52003397 | T | A | 0.02 |
| rs854037 | chr5:57091783 | A | G | 0.02 |
| rs28365970 | chr5:67585723 | C | A | 0.015 |

| SNP | Chrom:position (hg19) | Trait-raising Allele | Trait-lowering Allele | Beta (SEM[a] adjusted fetal effects) |
|---|---|---|---|---|
| rs1981627 | chr5:133838180 | G | A | 0.007 |
| rs2946179 | chr5:157886627 | T | C | 0.004 |
| rs34471628 | chr5:172196752 | G | A | 0.014 |
| rs9379084 | chr6:7231843 | G | A | 0.004 |
| rs35261542 | chr6:20675792 | C | A | 0.049 |
| rs9379832 | chr6:26186200 | A | G | 0.019 |
| rs9366778 | chr6:31269173 | G | A | 0.014 |
| rs6911024 | chr6:31368451 | C | T | 0.002 |
| rs9267812 | chr6:32128394 | T | C | 0.015 |
| rs1547669 | chr6:33775641 | G | A | 0.018 |
| rs75104038 | chr6:34190104 | A | G | 0.024 |
| rs9348981 | chr6:35687249 | T | G | 0.015 |
| rs7744700 | chr6:53349401 | T | A | 0.018 |
| rs76094073 | chr6:109288036 | G | C | 0.011 |
| rs6925689 | chr6:126865884 | T | C | 0.018 |
| rs6569647 | chr6:130337266 | T | C | 0.014 |
| rs6930558 | chr6:141878920 | T | G | 0.022 |
| rs962554 | chr6:142734204 | T | C | 0.015 |
| rs10872678 | chr6:152039964 | T | C | 0.028 |
| rs2934844 | chr6:166142456 | T | A | 0.018 |
| rs4719648 | chr7:2756832 | C | T | 0.014 |
| rs59084784 | chr7:22739562 | A | C | 0.011 |
| rs7808457 | chr7:22798265 | A | T | 0.002 |
| rs34776209 | chr7:23513093 | C | T | 0.015 |
| rs2908279 | chr7:44174857 | T | G | 0.007 |
| rs2971669 | chr7:44231778 | C | T | 0.003 |
| rs138715366 | chr7:44246271 | C | T | 0.235 |
| rs10265133 | chr7:45895604 | T | G | 0.02 |
| rs11983722 | chr7:46298647 | A | T | 0.029 |
| rs10265057 | chr7:47275737 | G | A | 0.036 |
| rs2237467 | chr7:50733316 | A | G | 0.011 |
| rs112139215 | chr7:73034559 | A | C | 0.056 |
| rs2282978 | chr7:92264410 | C | T | 0.021 |
| rs45446698 | chr7:99332948 | T | G | 0.017 |
| rs6467157 | chr7:127660763 | T | C | 0.014 |
| rs3918226 | chr7:150690176 | T | C | 0.005 |
| rs62496903 | chr8:6446938 | T | C | 0.028 |
| rs732563 | chr8:23345526 | C | T | 0.019 |
| rs11778247 | chr8:23403378 | A | G | 0 |
| rs34036147 | chr8:38366249 | T | C | 0.019 |
| rs13266210 | chr8:41533514 | A | G | 0.03 |
| rs72656010 | chr8:57122215 | T | C | 0.026 |
| rs6995390 | chr8:77611012 | A | T | 0.014 |
| rs7819593 | chr8:106115172 | C | T | 0.023 |
| rs10283100 | chr8:120596023 | G | A | 0.033 |

| SNP | Chrom:position (hg19) | Trait-raising Allele | Trait-lowering Allele | Beta (SEM[a] adjusted fetal effects) |
|---|---|---|---|---|
| rs13271368 | chr8:126506140 | C | T | 0.021 |
| rs13257363 | chr8:142252580 | G | A | 0.017 |
| rs9657468 | chr8:142362391 | G | T | 0.018 |
| rs7854962 | chr9:96900505 | C | G | 0.016 |
| rs28457693 | chr9:98217348 | G | A | 0.04 |
| rs2418135 | chr9:113901309 | A | G | 0.012 |
| rs72760655 | chr9:116916214 | A | C | 0.009 |
| rs1323438 | chr9:119115531 | C | T | 0.02 |
| rs3933326 | chr9:123633948 | G | A | 0.023 |
| rs10985827 | chr9:125701608 | G | T | 0.027 |
| rs28505901 | chr9:139241030 | A | G | 0.024 |
| rs4350272 | chr10:25056118 | A | G | 0.017 |
| rs9645500 | chr10:70986723 | G | T | 0.019 |
| rs1112718 | chr10:94479107 | G | A | 0.036 |
| rs10509669 | chr10:95969913 | T | A | 0.02 |
| rs3740360 | chr10:96025491 | C | A | 0.003 |
| rs2274224 | chr10:96039597 | C | G | 0.019 |
| rs562974282 | chr10:104201070 | T | G | 0.126 |
| rs10883846 | chr10:104958244 | C | T | 0.016 |
| rs7903146 | chr10:114758349 | T | C | 0.003 |
| rs7076938 | chr10:115789375 | T | C | 0.029 |
| rs71486610 | chr10:124134803 | C | G | 0.016 |
| rs11042596 | chr11:2118860 | T | G | 0.027 |
| rs234864 | chr11:2857297 | A | G | 0.017 |
| rs2168101 | chr11:8255408 | A | C | 0.015 |
| rs4444073 | chr11:10331664 | A | C | 0.023 |
| rs5030317 | chr11:32410337 | C | G | 0.007 |
| rs10437653 | chr11:46297631 | A | C | 0.002 |
| rs10734564 | chr11:48160429 | G | A | 0.009 |
| rs667515 | chr11:69449076 | G | C | 0.013 |
| rs61885091 | chr11:69791952 | A | G | 0.024 |
| rs10830963 | chr11:92708710 | C | G | 0.002 |
| rs10895278 | chr11:102095335 | T | C | 0.001 |
| rs76895963 | chr12:4384844 | G | T | 0.051 |
| rs11055030 | chr12:12878349 | G | C | 0.022 |
| rs2306547 | chr12:26877885 | C | T | 0.016 |
| rs11051061 | chr12:30914668 | A | G | 0.001 |
| rs6582623 | chr12:46613394 | C | T | 0.02 |
| rs180438 | chr12:47187260 | A | G | 0.007 |
| rs7968682 | chr12:66371880 | G | T | 0.037 |
| rs1480470 | chr12:66412130 | G | A | 0.028 |
| rs1533688 | chr12:102772745 | T | C | 0.004 |
| rs2647873 | chr12:103081192 | A | G | 0.009 |
| rs17033114 | chr12:103123339 | C | T | 0.008 |
| rs3184504 | chr12:111884608 | C | T | 0.005 |

| SNP | Chrom:position (hg19) | Trait-raising Allele | Trait-lowering Allele | Beta (SEM[a] adjusted fetal effects) |
|---|---|---|---|---|
| rs9549046 | chr13:40647206 | A | G | 0.027 |
| rs34217484 | chr13:48854550 | A | T | 0.012 |
| rs9318511 | chr13:78601413 | C | A | 0.024 |
| rs72681869 | chr14:50655357 | C | G | 0.108 |
| rs6575803 | chr14:101257755 | C | T | 0.034 |
| rs75844534 | chr15:38667117 | A | C | 0.036 |
| rs2928148 | chr15:41401550 | G | A | 0.004 |
| rs339969 | chr15:60883281 | A | C | 0.011 |
| rs3784789 | chr15:75082552 | C | G | 0.018 |
| rs12909648 | chr15:86224570 | A | G | 0.003 |
| rs12443252 | chr15:91064690 | C | T | 0.007 |
| rs4932373 | chr15:91429287 | A | C | 0.01 |
| rs55958435 | chr15:96852638 | A | G | 0.022 |
| rs7402983 | chr15:99193276 | A | C | 0.027 |
| rs11630479 | chr15:99240481 | G | A | 0.007 |
| rs2045457 | chr16:20046115 | G | A | 0.012 |
| rs40434 | chr16:55699525 | G | A | 0.017 |
| rs28544888 | chr16:55741204 | C | T | 0.027 |
| rs11641308 | chr16:75312023 | C | T | 0.005 |
| rs222857 | chr17:7164563 | T | C | 0.026 |
| rs4511593 | chr17:7455536 | T | C | 0.019 |
| rs78378222 | chr17:7571752 | G | T | 0.058 |
| rs9909342 | chr17:25652275 | A | G | 0.019 |
| rs7223535 | chr17:29211667 | G | A | 0.02 |
| rs11867479 | chr17:68090207 | T | C | 0.018 |
| rs10221267 | chr17:68464662 | T | C | 0.018 |
| rs73354194 | chr17:79905947 | C | T | 0.06 |
| rs9912553 | chr17:79959703 | G | C | 0.006 |
| rs11082304 | chr18:20720973 | T | G | 0.013 |
| rs2779165 | chr19:4915447 | G | C | 0.018 |
| rs8106042 | chr19:7161849 | G | C | 0.023 |
| rs2967676 | chr19:8789666 | C | A | 0.003 |
| rs41355649 | chr19:33790556 | G | A | 0.042 |
| rs1129156 | chr19:40719076 | T | C | 0.022 |
| rs147957154 | chr19:43431040 | T | C | 0.026 |
| rs516246 | chr19:49206172 | C | T | 0.017 |
| rs255773 | chr19:54723546 | C | T | 0.018 |
| rs147110934 | chr19:55993436 | G | T | 0.055 |
| rs12461110 | chr19:56320663 | G | A | 0.005 |
| rs304001 | chr19:56423668 | A | G | 0.003 |
| rs6040076 | chr20:10658882 | C | G | 0.015 |
| rs6033062 | chr20:11207419 | A | T | 0.014 |
| rs1203876 | chr20:22540915 | C | A | 0.055 |
| rs11698914 | chr20:31327144 | C | G | 0.029 |
| rs181451002 | chr20:32466219 | A | G | 0.006 |

| SNP | Chrom:position (hg19) | Trait-raising Allele | Trait-lowering Allele | Beta (SEM[a] adjusted fetal effects) |
|---|---|---|---|---|
| rs2889874 | chr20:33715777 | G | T | 0.014 |
| rs1012167 | chr20:39159119 | C | T | 0.024 |
| rs753381 | chr20:39797465 | T | C | 0.018 |
| rs6026449 | chr20:57272617 | C | T | 0.018 |
| rs73143584 | chr20:62445702 | A | G | 0.031 |
| rs2229742 | chr21:16339172 | G | C | 0.028 |
| rs220193 | chr21:43581308 | A | G | 0.018 |
| rs134594 | chr22:29468456 | C | T | 0.022 |
| rs41311445 | chr22:42070374 | A | C | 0.034 |
| rs7285579 | chr22:46441980 | C | T | 0.018 |

[a]SEM = structural equation model for partitioning of maternal and fetal effects. Note: genetic scores were unweighted i.e. all SNP weights set to 1 for analysis
